# Socioeconomic Status and Women’s Mental Health and Wellbeing in Male-Factor Infertility Marital Circumstances: A Scoping Review

**DOI:** 10.1101/2022.10.11.22280950

**Authors:** Nanji R. Umoh

## Abstract

**Background:** Infertility is established where frequent intercourse and non-contraceptive use over 12 to 24-month periods do not result in live births. It is an underemphasized global public health challenge occurring mostly in countries with the highest fertility rates. Infertility can be female- or male-factor-based, combined, unexplained, or impaired. Fertility research emphasizes socio-cultural beliefs, patriarchy, insufficient distinctions between masculinity and virility, etc., as impacting women’s mental health and wellbeing. Conversely, research, policies, and interventions underemphasize the male-factor as central to reproduction. This is counter-productive for their female counterparts in the reproductive equation and contributes to gaps in reproductive health literature. Male-factor infertility, a sensitive, long-neglected public health issue caused by genetic and environmental factors, constitutes 20% of infertility cases globally. The biopsychosocial impacts of childlessness on women in male-factor infertility circumstances are almost equally deleterious across social groups and regions.

**Objective:** To present a scoping review of evidence on the extent to which reproductive health literature recognizes socioeconomic status as central to women’s mental health and wellbeing in male-factor infertility circumstances.

**Methods:** Literature was mapped across five databases (MedlinePlus, Google Scholar, PubMed, ScienceDirect and Publons Web of Science) without restrictions to geographical regions. The Preferred Reporting Items for Systematic reviews and Meta-Analyses extension for Scoping Reviews (PRISMA-ScR) Checklist-2018 was the review protocol.

**Results:** 12 out of 2582 screened articles met the eligibility criteria. The impacts of childlessness on women in male-factor infertility circumstances are similar across regions, but aggravated by socioeconomic circumstances, particularly in low- and middle-income countries (LMICs) and societies that emphasize childbearing. Women with relatively high socioeconomic statuses and access to New/Assisted Reproduction Technologies (NRTs/ARTs) enjoy better mental health and wellbeing.

**Discussion/Conclusion:** The Social Determinants of Health (SDHs) provided the analytical framework. The socioeconomic status influences the quality of the women’s overall wellbeing in male-factor infertility circumstances, with implications for access to and affordability of New/Assisted Reproduction Technologies (NRTs/ARTs) and other related male-factor infertility treatment options. The deleterious impacts are more for those who are unable to afford the fertility treatments. Asides this socioeconomic context of infertility, a politico-legal context exacerbates the suffering of women in male-factor infertility circumstances, through policy gaps that exist in the provision of interventions to cater to the needs of socioeconomically disadvantaged women.

## Introduction

Infertility is a global health problem for which universal access to reproductive healthcare was instituted as a United Nations (UN) Millennium Developmental Goal (MDG) for 2015.^1^ It is currently subsumed in the Sustainable Development Goal (SDG-3) of Good health and well-being.^2^ Infertility describes the inability of couples to conceive despite non-contraceptive use^3^ and frequent, unprotected sexual intercourse over a one-year period,^4^ within the fertile phase of the maternal cycle.^5^ The World Health Organization (WHO) and European Society of Human Reproduction and Embryology (ESHRE) provide for a two-year minimum allowable period before a couple can be described as infertile.^6,7^ Infertility is disruptive,^8^ affecting aspects of men and women’s lives,^9^ and challenging the essential rights of individuals to the highest attainable levels of physical and mental health.^10^ It causes a temporary life crisis in most couples desiring to have children.^11^ It results in psychosocial impairments^12^ and stressful conditions which adversely affect marital relationships, sexual satisfaction,^11,13^ and equals suffering from cancer, hypertension and cardiac rehabilitation.^14^

The estimated global average prevalence of infertility is 9% among reproductive couples.^15^ 8-12% of couples around the world experience infertility at some point in their lives, bringing the number of sufferers to an estimated 48-186 million people worldwide.^16-18^ In America, it affects 10% of couples,^19^ and in the resource-poor regions of the world, an estimated 30% or more, given inconsistencies in defining infertility, and difficulties in data collation.^11,20-22^ Among the total infertile population globally, 40% is female factor-based, 30% combined male and female, 20% male factor-based, and 10–17% impaired.^23,24^ Infertility received comparatively little attention from social science researchers, anthropologists and demographers and was not portrayed as a serious problem.^22,25-28^ Most research and interventions by governments, agencies and development partners are directed at communicable diseases, fertility behavior,^29^ reduction of high fertility rates to ensure population control, and economic growth^30,31^ in LMICs.

National surveys show that infertility prevalence in Africa is higher than the rest of the world.^32^ Paradoxically, the highest rates of infertility are recorded in countries with the highest fertility rates.^5^ The plausible explanation is that the infertility rates are proportionate to the overall populations of given countries. Infertility in Sub-Saharan Africa ranges from 20-40%, yet the reproductive health strategies of countries in the region do not emphasize the reduction of its prevalence or impact.^33^ In the continent’s infertility belt stretching from West Africa, through Central, and to East Africa, records of high infertility prevalence^34-36^ constitute distinct and complex social and public health problems.^22,37^ Within the LMICs, most infertility results primarily from infectious diseases that damage the reproductive tract and secondarily from the unavailability or lack of access to fertility services.^11^

## Male-Factor Infertility

Male-factor infertility is the sole or contributing factor to infertility situations in a substantial proportion of involuntarily childless couples,^8,18,38,39^ making non-fatherhood more common than non-motherhood.^40^ Despite this, men are the ‘second sex’ in reproductive health research,^41^ historically overlooked due to misconceptions on their roles as elementary in human reproduction, whereby the male sexual organ is likened to a ‘mechanical instrument’ which either ‘works’ or does not.^42-44^ This marginalization of men is an oversight of considerable proportions^45^ that flows from the traditional assumption that a couples’ failure to conceive is the woman’s ‘fault’.^42,43^

Male-factor infertility describes the inability of a man to impregnate a woman after 12 months of regular, unprotected sexual intercourse.^4^ It is multifactorial. The rising morbidity rates are attributable to the deterioration of male sperm quality on an average of one percent annually, and a significant increase in sperm morphology and abnormality, globally.^46-49^ There are indicated associations between male infertility and exposures to sexually transmitted diseases (STDs), unorthodox (native) medication, and moderate to heavy alcohol consumption.^50^ Repeated episodes of penile discharge, painful urination, genital ulcers, erectile dysfunction, and testicular pain also directly affected their reproductive tracts. Other factors are genetics, physical and hormonal abnormalities, injuries, drugs, infections of the genital tract, infantile infections, exposure to environmental factors including diet, radiation and toxic elements, cultural behaviors, otherwise unexplainable causes,^43,44,51,52^ or idiopathic factors.^53^ The damage by these factors is sometimes irreversible.^5^

## Objective

The research explores the socioeconomic context of infertility by mapping reproductive health literature on women in male-factor infertility marital relationships, to determine whether their socioeconomic status is adequately considered as a factor in their wellbeing in such circumstances.

## Rationale

The male-factor infertility diagnosis diminishes couples hopes for having a baby, and typically portends common problems for the women. Women are considered as primary in reproduction because it is they who become pregnant.^87,^ As such, most fertility problems and diagnostic and treatment regimes are typically assumed to affect them,^37,87^ thereby making the male-factor infertility of less significance.^61^

This overemphasis on fertility as a female reproductive health issue portends dire consequences for the ‘barren’ women. They are usually the most traumatized, disenfranchised and disinherited as a result of their inability to have children, even in proven male-factor infertility marital relationships, and especially in societies with emphasis on childbearing. It leads to feelings of failure, incompleteness and uncertainty as women, lack of sexual identities, hopelessness and despair, physical and psychological stress from forms of violent and non-violent abuse, and sometimes depression, because of the inability to fulfil the childbearing obligations.^88^ A study by Savadzadeh, et al showed that infertility results in destructive emotional experiences, feelings of insecurity, and worry about its effects on lived experiences and on the stability of marital life.^89^

The majority of research on the impact of male-factor infertility on women’s mental health reflect these perspectives as the trajectory of women’s suffering in infertility circumstances. This is particularly so because of the social, economic and community determinants which make being able to bear children a very important aspect of societies, and more especially, those with emphasis on childbearing.^43,90^ This research therefore explores the possibility of better wellbeing outcomes that relate to the women’s socioeconomic statuses, and the access to or affordability of fertility treatment options that enable them bear children.

## Conceptual Framework: The Social Determinants of Health (SDHs)

The SDHs framework defines health as determined by the social conditions in which people are born, grow, live, work and age; as well as the impact of the inequitable distribution of power, resources, and the policy directions and actions^54^ on health equity in society. The structural and intermediary determinants include the socioeconomic and political contexts, the social positions, the healthcare system, and the distribution of health and wellbeing that influence the consequences of illness in people’s lives.^54^

‘Context’ defines the sociopolitical mechanisms of policy, governance, socio-cultural values, occupation, income, gender, education, race/ethnicity, social cohesion, social class, and the behavioral, psychosocial and biological factors that combine to determine quality of life.^54^ Poland, et al highlight the importance of social contexts of behaviour as consequential to understanding the broader social determinants of health. Depending on the society to which they are applied,^55^ contexts form the environment within which something exists, occurs, and phenomena become intelligible and meaningful.^54^

### The Social and Cultural Contexts of Infertility

Local levels, values, trends and socio-demographic patterns of infertility require an understanding of the meanings and consequences for individuals.^22^ In the LMICs, the experience of infertility is shaped by culture, patriarchy and religion.^8,19,56^ Cultures prioritize fertility and motherhood as central to women’s identities, their achieving adult status and acceptance in their communities.^13,22^ In Zimbabwe, the birth of children gives a woman the right to share in her husband’s property and wealth.^57^ In Yoruba culture, the adult woman’s role depends on motherhood because children ensure the continuation of lineages.^57^ In Cameroon, Nigeria and some Asian countries, infertility is a source of poverty for women.^57^ In Egypt, women bear the burden of infertility even when there is a known male cause, because having a child completes the woman more than it does a man. Throughout all social classes, the belief is that fertile or not, a man is always a man.^8^

In Bangladeshi slums the ‘treatment’ for males is remarriage, as women are held responsible for infertility.^57^ In Costa Rica, having children is a natural, uncontrollable process embedded in God’s will. Infertile couples are fatalistic in their perceptions rather choosing to adapt to the undesirable fate. In a study, a woman resigned herself to childlessness when her husband refused to be tested.^58^ Sometimes, they explore spiritual and holistic measures including prayers, consulting healers and herbal remedies, to resolve the infertility, before contemplating orthodox medical solutions.^57,58^ In Rwanda, couples’ relationships are affected by gender-specific diagnoses. Infertile women were more likely to be abused or divorced than female partners of infertile men, while infertile men do not expect to be abandoned by their wives.^59^ Nevertheless, most women seek treatment for male-factor infertility to avoid upheavals in their personal or social life.^8,42,59^

In most LMICs, parenting is socio-culturally considered a core developmental milestone in adulthood.^44,60^ Societal and parental pressures for propagation of the family name psychologically affect infertile couples lives and identities.^13,61,62^ Infertility is regarded as a serious failure of the woman to fulfill her role.^14,61,62^ Some customs and traditions lead to negative attitudes towards ‘infertile’ women.^63^ They become care-givers for the sick and infirm,^64,65^ or for others’ children,^64,66^ and are not allowed to inherit or continue living in their husbands’ compounds after their deaths.^22,44^

Some men’s personal experiences of reproduction are mediated by numerous biopsychosocial systems of meaning attached to their masculinity and virility.^13,37^ Many infertile men rather suffer from low self-esteem, anxiety, isolation, blame, and greater sexual inadequacy^57^ than respond to pain and illness by seeking healthcare.^67^ Most times, wives seek medical help before their husbands, enduring blames and the fertility treatment routines for the childlessness.^61,68-70^ Consequently, they develop more distress in response to the social constructs and outdated gender stereotyping that is unsupported by research data.^71-73^

In African cultures, childbearing relates with the individual’s identity, and perceptions of self-perpetuation and mortality.^74,75^ In the traditional Chinese society, fertility and procreation are influenced by social and cultural contexts^44,76^ within which childlessness is treated as a dishonor to parents.^57^ In African, Middle-East and Asian societies, the presence of children allows their parents, mothers especially, to be better esteemed members of the family and community.^77^ In rural, less privileged areas, children are a reliable source of manpower^22,78-80^ and economic security especially in old age.

In Nigeria, Egypt, Iran, and China, male-factor infertility is responsible for about 50% of all infertility cases,^4,8,19,44,61^ yet social and cultural values divert the spotlight from the men whenever it occurs in a marital relationship.^43^ Some couples deal with it as a closely-guarded secret, to protect the ‘masculinity’ of the husbands,^8^ and little is done to address the infertility. To cope, some Nigerian women engage in extramarital affairs to be impregnated by other men, either to prove their ‘innocence’ or to save their husbands faces. Others endure name-calling, taunts, etc.,^43^ or get divorced and remarry, especially in Islamic societies where polygamy is acceptable.^8,13^ In some LMICs, the rights and interests of the man, the family and the community supersede those of the woman whose status is considered a derivative, sometimes inconsequential one.^81^ This inhibits women from asserting themselves when there is male-factor infertility.

In Iran, the family is an open system where couples are influenced by their own families and relatives. These influences impose psychological and societal burdens, and responsibility for pregnancy on the women than men, exposing most infertile people to undesirable reactions within their social environments.^61,82-85^ Consequently, the pain of coping with the infertility is aggravated above the infertility and its treatment, into a pain associated with society.^85^ In developed countries, there have been demographic transitions whereby infertility is sometimes voluntary due to the ‘choice’ factor that allows women exercise the right not to have children.^86^

## Methods

The Preferred Reporting Items for Systematic reviews and Meta-Analyses extension for Scoping Reviews (PRISMA-ScR) Checklist-2018^91^ provided the layout for this review, with additional guidance drawn from the Arksey and O’Malley framework.^92^ The review of literature across the databases (ScienceDirect, Google Scholar, PubMed, MedlinePlus, Publons Web of Science), was to enhance the validity and reliability of the study, enable a holistic appreciation of the phenomenon,^93^ and ensure the reproducibility of the study. It turned up an array of articles on the social, psychological, economic, cultural and physical impacts of infertility on couples. The review process was guided by the dominant themes - socioeconomic status, male-factor infertility and women’s mental health. Socioeconomic status was defined as appertaining income, affordance of infertility treatments, and occupational and educational levels.

**Figure 1:**
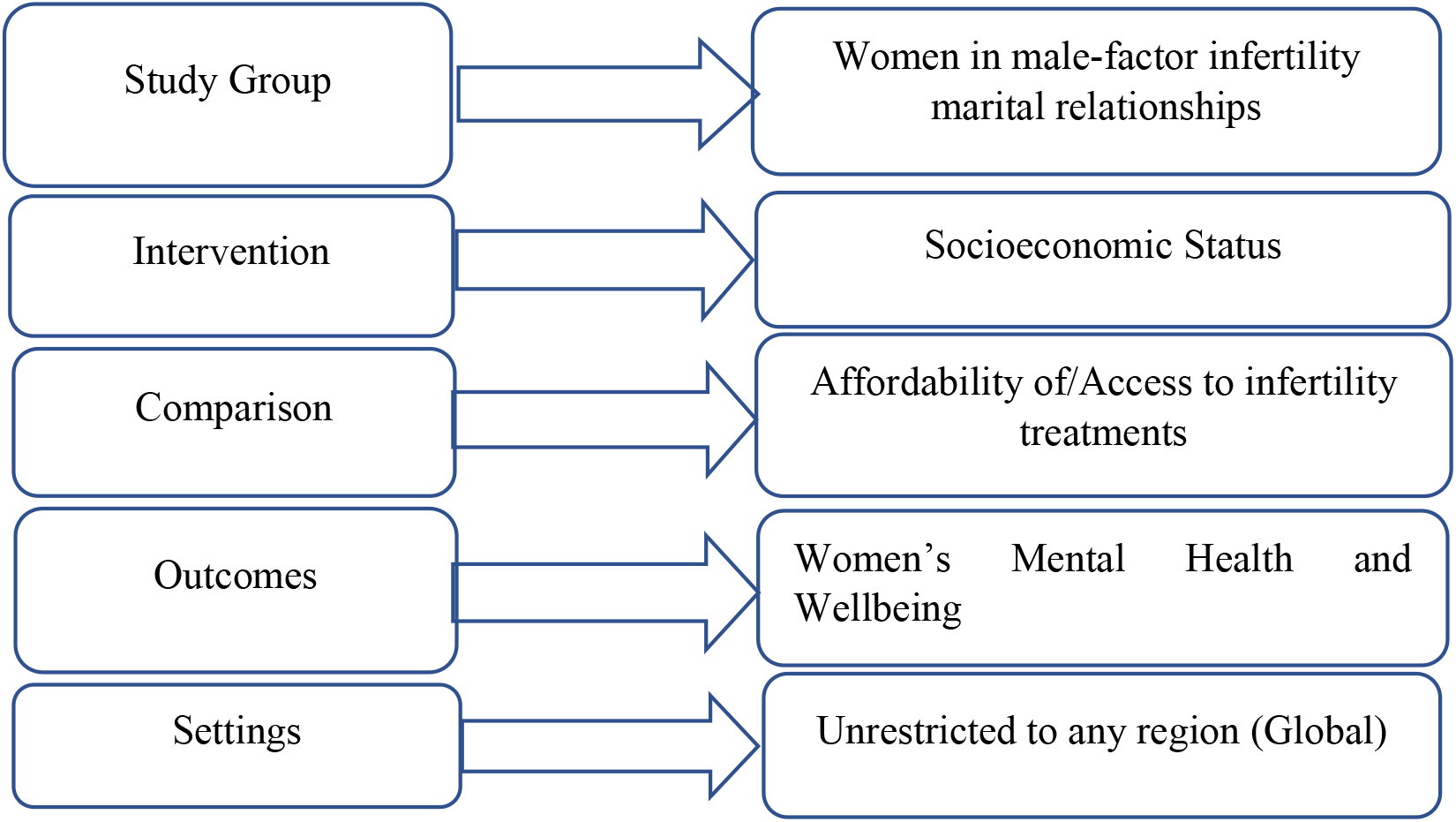
Variables, Settings, Populations, and Outcomes.

The study comprised women in male-factor infertility marital relationships. The intervention variable is the women’s socioeconomic status. The comparison factor explains the women’s mental health outcomes in relation to access to and affordability or otherwise, of fertility treatments.

### Inclusion Criteria

Included studies were:

- Alluding to women’s improved mental health, wellbeing, and coping abilities from access to/affordability of fertility treatments, or trauma from non-affordability.
- Published between 1997 and 2021.
- Published in English language.

### Exclusion Criteria

Studies with the following characteristics were excluded:

- Female-factor infertility in marriage;
- Not discussing the socioeconomic status of the women as impacting their mental health in male-factor infertility marriages;
- Impact of male-factor infertility on men’s mental health
- Not incorporating socioeconomic status, male infertility and women’s mental health;
- Infertility studies in non-human subjects.

There was no specificity in regard to the type, severity or otherwise of the male-factor infertility. The socioeconomic status is explained as an enabler of women’s access to fertility treatments in male-factor infertility circumstances. Since there was no formal assessment of bias, the inclusion criteria were expanded to reduce the risks of selection bias.^93^

### Search Strategy

Combinations of search terms on MedlinePlus, Google Scholar, ScienceDirect, and PubMed turned up an intangible number of articles, too few to ensure the reliability of the results. Subsequently, one set - Socioeconomic Status, Male Infertility, Women’s Mental Health - was utilized across all the databases since it yielded a considerable number of articles after an initial test-run on Publons Web of Science. Searches of studies between 2010-2021 did not yield sufficient information on the three main variables. Findings showed that studies in andrology and male reproductive health in the LMICs gained grounds around the mid-1990s. Between 2005 and 2021, NARTs became more acceptable in the LMICs. Stigmatization was greatly reduced, allowing women who were able to afford the services, subscribe to it.^50^ To enhance the currency of the research, the scope of the search was broadened to 1997-2021. Article quality and geographical locations were not criteria for inclusion or exclusion.

**Table 1:**
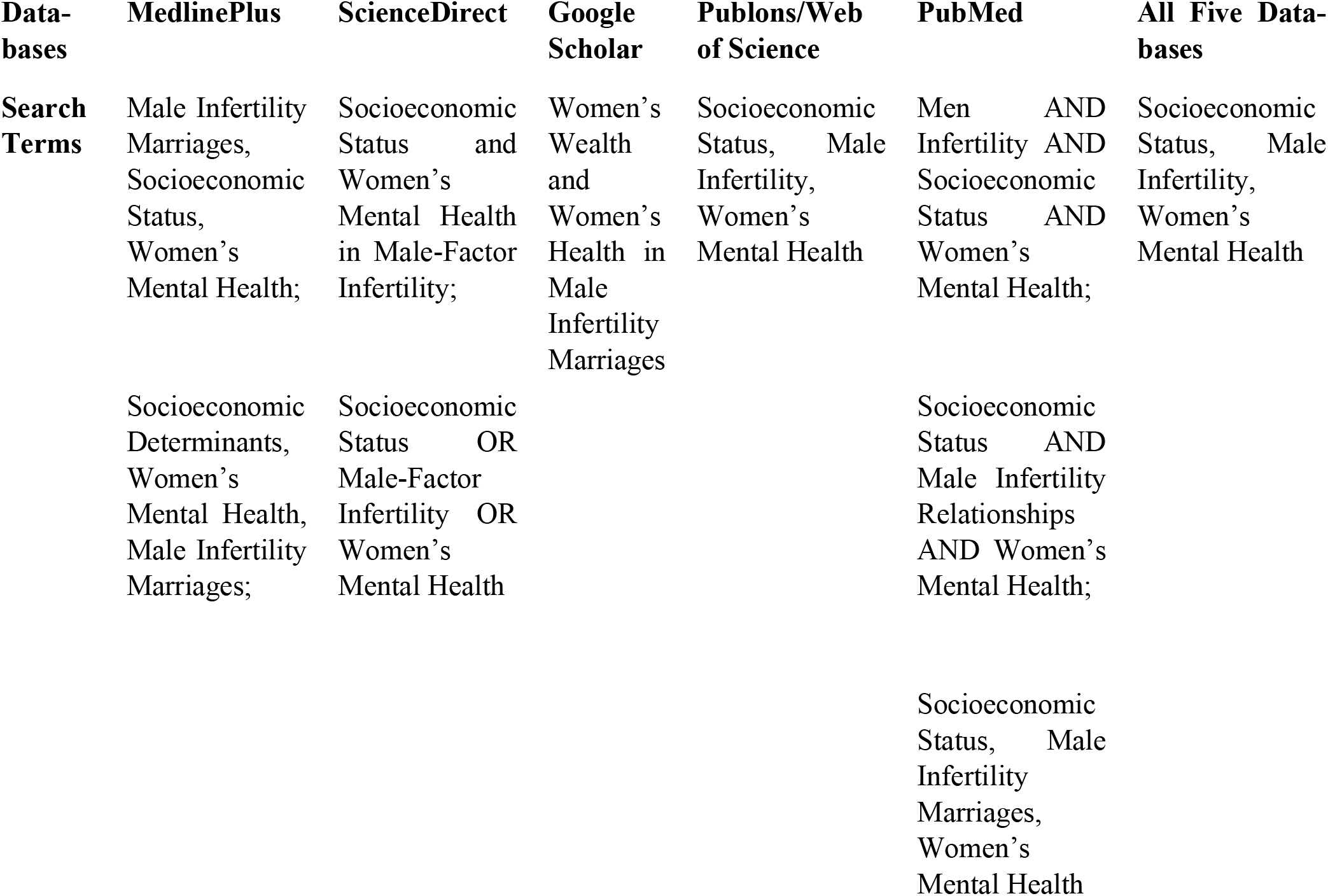
Search Term Combinations and Databases.

| **Data-bases** | **MedlinePlus** | **ScienceDirect** | **Google Scholar** | **Publons/Web of Science** | **PubMed** | **All Five Data-bases** |
| --- | --- | --- | --- | --- | --- | --- |
| **Search Terms** | Male Infertility Marriages, Socioeconomic Status, Women's Mental Health; | Socioeconomic Status and Women's Mental Health in Male-Factor Infertility; | Women's Wealth and Women's Health in Male Infertility Marriages | Socioeconomic Status, Male Infertility, Women's Mental Health | Men AND Infertility AND Socioeconomic Status AND Women's Mental Health; | Socioeconomic Status, Male Infertility, Women's Mental Health |
|  | Socioeconomic Determinants, Women's Mental Health, Male Infertility Marriages; | Socioeconomic Status OR Male-Factor Infertility OR Women's Mental Health |  |  | Socioeconomic Status AND Male Infertility Relationships AND Women's Mental Health; |  |
|  |  |  |  |  | Socioeconomic Status, Male Infertility Marriages, Women's Mental Health |  |

**Figure 2:**
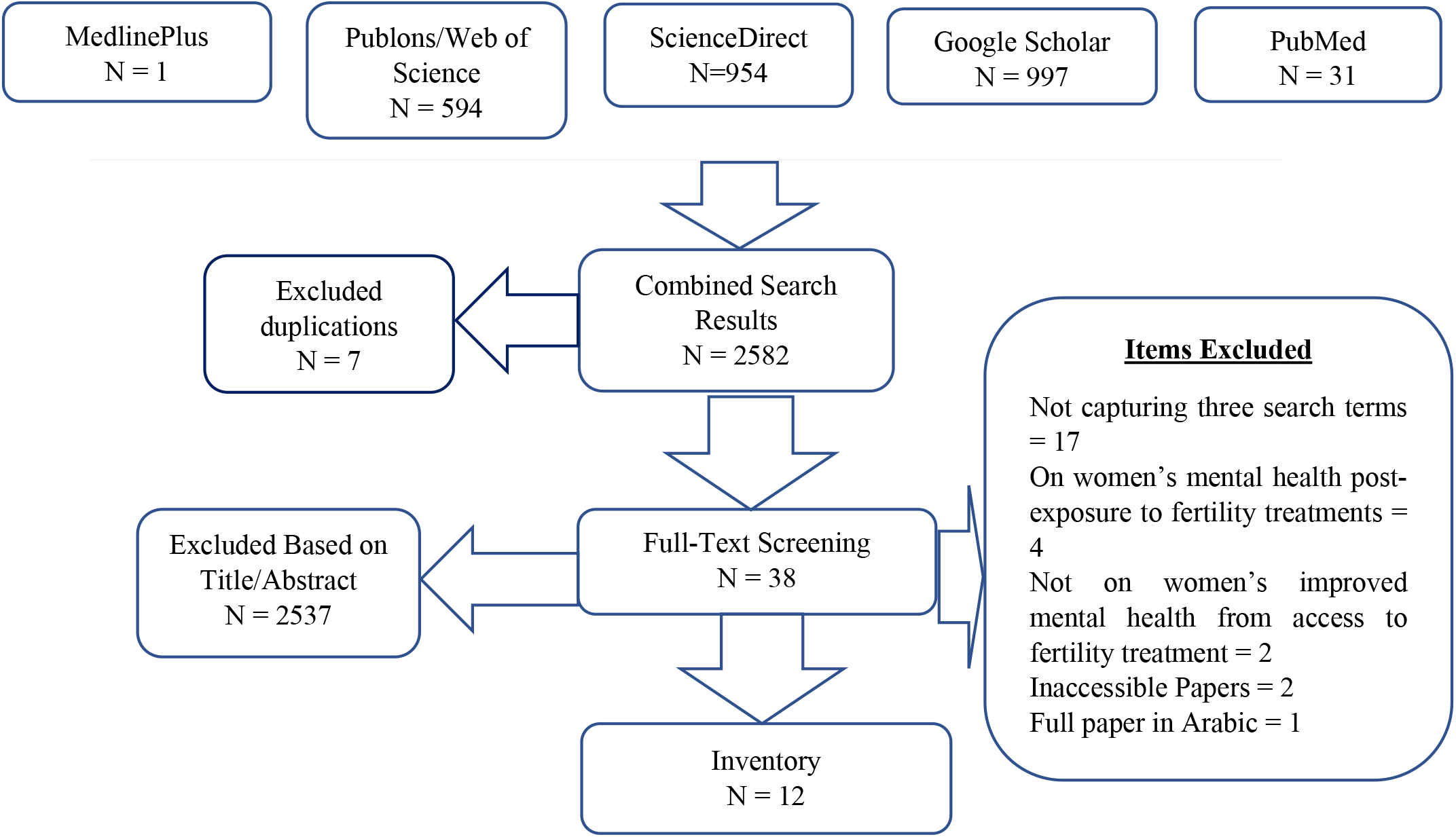
Database Summary.

The searches returned 2,582 results which were subjected to further screening. Seven duplicate articles were removed. 2,537 articles were excluded through the titles and abstracts (TIAB) screening. 38 articles were eligible for full-text screening.

Of the 38 articles, the full-texts of three were inaccessible. Two were not open access, the full-text of the third was in Arabic, despite its abstract in English. 17 papers did not capture the three main themes - socioeconomic status, male infertility, women’s mental health. Four articles were focused on the impact of NARTs on the women’s mental health. One paper was focused on coping strategies in male-factor infertility marriages and the last paper, on the effect of male-factor infertility on women’s sexual satisfaction. Consequently, based on full-text screening, only 12 papers were eligible for inclusion in the scoping review.

### Data Charting

Data on the authors, title, year of publication, journal, study design, sample size, methods of data analysis, participants demographics, study durations, settings, and outcomes were extracted from each article and presented on a table.

**Table 2:** Data Extraction Summary.

| DATA EXTRACTION/SUMMARY TABLE |  |  |  |  |  |  |  |  |  |  |  |  |  |  |
| --- | --- | --- | --- | --- | --- | --- | --- | --- | --- | --- | --- | --- | --- | --- |
|  | AUTHOR(S) | PAPER TITLE | YEAR OF PUBLICATION | JOURNAL | STUDY DESIGN | DATA COLLECTION AND ANALYSIS METHODS | PARTICIPANTS |  |  |  |  | STUDY DURATION | SETTING | OUTCOMES |
|  |  |  |  |  |  |  | COUNTRY | AGE | SEX | DIAGNOSIS | SAMPLE SIZE |  |  |  |
| 1 | Bai, et al. | Gender differences in factors associated with depression in infertility patients | 2019 | J Adv Nurs. | Cross-Sectional Study | Questionnaires, Descriptive Statistics | CHINA | Mean Ages - Men: 31.08, Women: 28.84 | 380 women, 360 men | Female and Male Infertility | 740 | Unspecified | Rural, Suburban, Urban | There is infertility-related stress and depression from the economic burdens related to cost of fertility treatments |
| 2 | Baybordi, E. | The role of social determinant of health in couples' infertility | 2019 | Social Determinant of Health and Infertility | Descriptive Cross-Sectional Study | Questionnaires, Descriptive Statistics | IRAN | Mean Age - 33.78 | 66 Male, 48 Female, 54 Couple | Female, Male and Couple Infertility | 205 | Six Months | Infertility Treatment Centre | There is a prevalence of male-factor infertility across all groups. Lower income and socioeconomic status resulted in lower quality of life |
| 3 | Greil, et al. | The Social Construction of Infertility | 2011 | Sociology Compass | Qualitative Study | Comparative Thematic Analysis | Cross-national | Unspecified | Unspecified | Female and Male Infertility | Unspecified | Unspecified |  | Infertility and reproduction are stratified social concepts. Those in higher classes are more advantaged with rights and choices, unlike those in the lower classes |
| 4 | Hammerberg, K. & Kirkman, M. | Infertility in resource-constrained settings: moving towards amelioration | 2012 | Reproductive BioMedicine Online | Qualitative Study | Comparative Thematic Analysis | Cross-national | Unspecified | Unspecified | Male and Female Infertility |  | Unspecified |  | Access to fertility treatment is inequitably related to income and socioeconomic status with dire consequences |
| 5 | Jahromi, et al | Quality of Life and Its Influencing Factors of Couples Referred to An Infertility Center in Shiraz, Iran | 2018 | Int J Fertil Steril. | Cross-Sectional Study | Interviews, Questionnaires, Content Analysis | IRAN | Mean Ages - Men: 32.30, Women: 31.66 | Male - 147, Female - 132, Both - 45, Unexplained - 130 | Male, Female, Both, Unexplained Infertility | 501 | 13 Months | Infertility Treatment Centre | Shorter durations of infertility and male etiology have higher Quality of life (QoL) for the women. Lower academic education and lower income levels restrict access to treatment options. Prior unsuccessful treatments are associated with low QoL |
| 6 | Inhorn, M. | The Worms Are Weak: Male Infertility and Patriarchal Paradoxes in Egypt | 2003 | Men and Masculinities | Qualitative Study | Interviews | EGYPT | Unspecified | Unspecified | Male Infertility | Unspecified | 13 Months with Urban poor, then 3 Months with Urban Middle and Upper middle classes | Urban Public Infertility Treatment Centre | Access to fertility treatment improves the women's quality of life but it is hampered by societal norms of patriarchy |
| 7 | Onat, G. & Beji | Effects of infertility on gender differences in marital relationship and quality of life: a case-control study of Turkish couples | 2012 | European Journal of Obstetrics & Gynecology and Reproductive Biology | Case-Control Study | Quantitative Methods | TURKEY | Unspecified | Male - 58, Female - 58 | Male and Female Infertility | 116 | 14 Months | Urban Public Infertility Treatment Centre | Couples who attended the clinics and sought treatment for infertility had strong marital relations |
| 8 | Schmidt, et al | The social epidemiology of coping with infertility | 2004 | Human Reproduction | Cross-Sectional Study | Questionnaires, Descriptive Statistics | DENMARK | Mean Ages - Men: 34.1, Women: 31.8 | Male - 1081, Female - 1169 | Male and Female Infertility | 47 - Private, 1406 Couples (2812 people) Public | 19 Months | Public and Private Fertility Clinics | Individuals (women and men) in the higher socioeconomic classes cope better with the infertility (male or female - as is relative) |
| 9 | Stevenson, et al | Through women's eyes: The female experience of infertility following a recent diagnosis of severe male factor infertility | 2019 | Human Reproduction | Qualitative Study | In-depth Interviews | US and UK | 40 years or younger | Female partners | Male Infertility | 11 | 28 Months | Urban Infertility Treatment Centre | The psychosocial impact of male-factor infertility is understood from the partner's perspective; Couples who attended the clinics and sought treatment had strong marital relations |
| 10 | Taghipour, et al | Women's perceptions and experiences of the challenges in the process of male infertility treatment: A qualitative study | 2017 | Electronic Physician | Qualitative Study | Interviews, Content Analysis | IRAN | Women's Mean Age - 34.27 | Female partners | Male Infertility | 30 | 12 Months | Urban Public Infertility Treatment Centre | Access to fertility treatment improves quality of life but is hampered by treatment related stress on the women partners |
| 11 | Bell, A. V. | Overcoming and (Maintaining) Reproductive Difference: Similarities in the Gendered Experience of Infertility | 2015 | Qual Sociol | Qualitative Study | Content Analysis | US | Mean Age - 35 | Male - 26, Female- 17 | Male and Female Infertility | 43 | Unspecified |  | Access to fertility treatment is stratified, improves quality of life but is hampered by societal norms that make infertility a women's issue. Infertility should be a couples' issue |
| 12 | Vizheh, et al. | Impact of Gender Infertility Diagnosis on Marital Relationship in Infertile Couples: A Couple Based Study | 2015 | Sex Disabil | Cross-Sectional Study | Questionnaires, Descriptive Statistics | IRAN | Mean Ages - Men: 32.1, Women: 27.13 | Male - 32, Female - 48, Both - 19, Unexplained - 24 | Male, Female, Both, Unexplained Infertility | 123 Couples | Unspecified | Urban Public Infertility Treatment Centre | In the male infertility instances, the women with higher educational statuses fared better than those with lower educational statuses |

## Results

12 articles met the inclusion criteria. 2/12 were cross-national comparative studies on male and female infertility in different countries. The other ten were spread across Iran (4/12), US (1/12), US and UK (1/12), Turkey (1/12), Egypt (1/12), China (1/12), and Denmark (1/12). 5/12 were cross-sectional studies conducted over three to six-month periods. 1/12 was a case-control study in Turkey and 6/12 were qualitative studies which utilized content analyses for data handling. 4/12 were focused solely on male infertility and the other variables of socioeconomic status and the mental health and wellbeing of their female spouses. 8/12 discussed male and female-factor infertility in marital relationships, socioeconomic status and the wellbeing of the spouses. However, only data appertaining to male-factor infertility, socioeconomic status and the mental health and wellbeing of their female spouses were extracted for this report.

### Socioeconomic Status and Women’s Mental Health in Male-factor Infertility Circumstances

The medicalization and stratification of infertility leads to comparisons between infertility help-seeking and reproductive behavior in developed and less developed societies.^57^ The development of fertility drugs evolved into new and assisted reproductive technologies (NARTs) such as in vitro fertilization (IVF) and intra-cytoplasmic sperm injection (ICSI) to address infertility.^21,57^ Stevenson, et al suggest that the psychosocial impact of male-factor infertility is better understood from the partner’s perspective.^60^ Women reported strong marital relations and positive relationship quality from access to NARTs.^60^ Onat and Beji also reported that the female partners in the male-factor infertility group fared better as a result of access to fertility treatments.^14^

From participants’ experiences, access to resources and fertility treatment is stratified, and specific to the populations with higher socioeconomic status.^94^ Women in higher socioeconomic groups are more advantaged than those in the lower social groups and can choose certain treatments unlike their counterparts in the lower socioeconomic groups.^57^ Affluent women in The Gambia, India, and Egypt are able to afford treatments and access sophisticated gynecological facilities and NARTs, while the needs of poor and middle-class women are not met.^57^

In many advanced nations, access is an issue for ethnic minorities and those with lower socioeconomic status.^57^ White men of high SES enact and define masculinity differently from the ways that men of racially and economically marginalized backgrounds do,^94^ yet the ability to detach fertility from virility through the medicalization of infertility option, is unavailable to men of low SES.^94^ In the United States, healthcare is based on a market model, making it one of the societies with the most exorbitant rates for fertility treatments.^59^ Most states do not mandate insurance coverage, and access to NARTs is inequitably related to income and socioeconomic status, raising infertility care beyond the reach of low-income groups.^57,59^

Statistics in the US reveal that women of low SES experience greater difficulty conceiving children, meanwhile, infertility is generally understood to be a wealthy, white woman’s issue. Historical stereotyping of women of lower SES and women of color, portray them as overly fertile, with US social policies further entrenching these ideas.^59^ ‘Stratified reproduction’ introduced by Colen and popularized by Ginsburg and Rapp^57^ describes the structuring of reproduction across social and cultural boundaries that empower privileged women, and disempower less privileged women to reproduce.^57^ Notions on stratification are applicable to the context of access to care which is severely limited in developing societies.^57,59^ Such depictions shape women’s infertility experiences beyond the medical, to the often overlooked economic and classist underpinnings of reproduction, family, motherhood, and health.^59^

Bai, et al discussed the impact of infertility on spouses in low-income groups in research-neglected, resource-poor areas in China.^44^ Infertility in China imposes heavy financial burdens on families because treatment costs entail full out-of-pocket payments accounting for more than 40% of annual non-food expenditure,^44^ resulting in catastrophic expenditures for infertile couples.^44^ Though women in male-factor infertility circumstances reported improved health from access to infertility treatment,^44^ with gender-role expectations, the men are worse-hit as the heads of homes, and the primary sources of most of the household income.^44^

In France and Israel, infertility treatments are subsidized, so the socioeconomic status does not significantly influence the utilization of ARTs.^95^ In Denmark where access to NARTs is free and offered in a tax-financed health system to ensure equal and easy access, focus is on the relationship between socioeconomic circumstances and coping with infertility.^95^

Highly educated people and people from higher social groups are less avoidant, engaging with active problem-focused strategies to figure out causes, assess situations, seek information, and express their emotions per the infertility problem.^95^ Couples from lower social groups cope through relationships with family, friends and colleagues with children than couples from higher social classes.^95^ Overall, the women are confronted with the infertility in all spheres of life, while the men engage in extra-curricular activities as ‘pain-free zones’ detached from their infertility stress.^95^

In Iran, Jahromi, et al found that in male infertility circumstances, low income and educational levels, lack of access to treatment and records of unsuccessful treatments, resulted in reduced quality of life (QoL) for women.^60^ The male-factor infertility couples with shorter durations of infertility experienced higher QoL.^60^ Iranian women with higher educational statuses fared better than those with lower educational statuses.^13^ Baybordi reported a prevalence of male-factor infertility across all social groups in Iran. The study also revealed that lower income and socioeconomic status resulted in lower QoL especially for the women.^19^ Most infertile people had undesirable income levels. The costs of NARTs made them even poorer, and more than other non-medical causes, exacerbated mental turbulence in the women spouses, accounting for couples’ distress.^19,61^

Social support for individuals in infertility circumstances is influenced by the income levels, though not significantly, due to the cultural status and the belief in family unity.^19^ The ‘ratio’ of social support received, is proportional to the socioeconomic status of the infertility sufferers.^19,61^ All participants in the lower-income groups stated that they enjoyed low to moderate social support, while individuals with moderate to high incomes enjoyed lots of social support and more unity from family and friends.^19,61^ Inhorn noted that access to fertility treatment improves QoL,^8,61^ but is hampered by societal norms of patriarchy that makes the stress of infertility less profound than the patriarchy itself because women prefer to keep their husbands secrets, than be divorced.^8^ However, some Egyptian women enjoyed loving relationships with their infertile husbands who were less likely to practice polygamy.^8^

## Discussion

### The Socioeconomic Context of Infertility

Infertility affects male and female, disproving perceptions of infertility as a women problem.^16^ The overemphasis on fertility as a female reproductive health issue portends dire consequences for the ‘barren’ women.^96^ Fertile women with infertile spouses experience similar infertility stress since regardless of the cause of infertility, the eventual outcome is childlessness.

The prevalence and impact of infertility are similar across low-, middle-, and high-income countries. The socioeconomic status of the women influences their mental health and wellbeing in male-factor infertility circumstances, yet is understudied in reproductive health discourses.^41-43,45,57,97^ The WHO reports that upper socioeconomic classes live better, longer lives, and suffer less from diseases than the poor.^98,99^ This confirms health inequality as a major social injustice underlying some of the strongest impacts on population health globally.^98^

At the micro level, the psychosocial, sociocultural, and economic consequences for low-income women with involuntary childlessness in LMICs, are more severe than in most Western societies. Access to infertility care is varied and usually only attainable by the wealthy,^1^ placing infertility among the most well-known diseases^19,100^ that require immediate interventions to cushion its effects on women’s mental health. In most patriarchal societies in the LMICs where the headship of the home rests on the man,^8,13,19,40,61,97,99^ the socioeconomic statuses of the women are socio-culturally determined by or related to those of their husbands.^8,44^ Sometimes this automatically gives him an edge over the female partner in the marriage especially where he has more financial dominance, and restricts her access to the finances.

The studies indicate that women in male-factor infertility situations are open to accessing the NARTs to improve their circumstances, but their socioeconomic and sociocultural realities limit their choices, capacities, and perceptions of themselves in marital circumstances. Alongside the women’s desires to have children, a culture-based commitment to the marital relationships downplays tendencies for divorce.^19,61^ Only in one instance in the Egypt study, was a divorce initiated by the infertile husband, on account of the deterioration of the marriage due to the childlessness, to maintain his ‘headship’ of the home.^8^

Couples greater socioeconomic resources have a stabilizing effect on their marital relations. Extensive resources provide material rewards that satisfy physical subsistence, safety and psychological security needs, and simultaneously offer symbolic rewards like higher social statuses. These ‘insure’ against divorce, making spouses reluctant to unsettle the assets of the family.^101^ There is an ‘income effect’ when the woman’s resources enhance the total resources of the family, and increase marital stability.^102^ Despite this possibility, the study findings neither referenced the women’s financial independence nor expressly discussed scenarios where the resources of the household are solely or highly dependent on the women. Rather, opting for the NARTs is tied to the ‘acceptance’, cooperation and sponsorship of their spouses. Financial difficulties and the man’s inability to fulfil his obligations as the primary breadwinner undermine mutual esteem and affection among partners, add to psychological distress, and contribute to interpersonal tensions.^103,104^ This lends credence to findings on the place of the socioeconomic status particularly in most LMICs whose social contexts are characterized by dire economic realities.^13,19,44,57,59,61,77^

From the results, a politico-legal dimension of the socioeconomic context of infertility within the LMICs and the developed countries exists. It provides a macro perspective on how the dearth of interventions allows the women’s socioeconomic status confine them within this category, and continue to impact their mental health and wellbeing in male-factor infertility circumstances. In these LMICs, there is a low priority for public funding, state-sponsored infertility care remains marginal, difficult to implement, and excludes access to or coverage of NART services.^105,106^ Fertility treatment cycles entail prohibitive out-of-pocket expenditure that costs more than half of an average individual’s annual income,^107^ and financially incapacitates individuals with scarce resources.^29,59^ The NARTs are available in the private sector, only accessible to the wealthy elite who can afford to pay.^29^

Few Western societies incorporate the diagnosis and treatment of infertility into their family planning programs.^59,95^ The criticism is that social equity-inclined policy actions are downplayed, invariably reducing the access of disadvantaged social groups to healthcare services.^108,109^ Across all regions, a systemic ‘structural violence’^110^ epitomized by the low attention of research, health planning, policy-making, and interventions^98,99^ to cushion the costs of access to fertility treatments for socioeconomically disadvantaged women with infertile spouses, exposes this group of women to myriads of infertility-related psychosocial health challenges.^61,94,111,112^ The lack and non-empowerment of women economically, allows them remain in vulnerable and disadvantaged positions, exposed to the impacts of health inequalities.

Rationales and policy designs for cross-government commitments to comprehensive reproductive healthcare management and cost-effective infertility services should accommodate the requisite SDHs affecting low-income groups,^59,98,99^ and be relative to the cultural realities of specific regions.^18^ Strategies for intervention packages must be tailored to peculiar contexts and exhibit an explicit understanding of how differences in SES affect intimacy. Access to fertility treatments, especially in the LMICs where cultures emphasize patriarchy and childbearing, and all forms of infertility are treated as women’s issues, is central. Policy makers should adequately plan for infertile couples to receive necessary and suitable economic, psychosocial, and medical support for increased access to the treatment modalities. The implementation processes must be tailored to ensure that fertility treatments are public goods that do not result in catastrophic or impoverishing out-of-pocket payments and outcomes.^44^

## Limitations

All available data on the study may not have been identified.^93^ Grey literature, relevant data outside the research timeframe, or inaccessible due to subscription, language, or other restrictions may have been left out. Data may have been lost through the TIAB screening process, where abstracts were inconsistent with the full-texts of the papers.^113^Scoping reviews are time-consuming, and do not provide synthesized results or answers to specific questions rather, they provide overviews of available literature on areas of research.^93^ This review highlighted gaps relating to dearth of literature on women’s socioeconomic status and implications for their mental health and wellbeing.

## Conclusion

The meanings of fertility and infertility are localized and specific, depending on the dominant perspectives held in the societies in which they occur. The combination of the social, cultural and economic values influences the social, socio-cultural, socioeconomic and politico-legal contexts of infertility that determine the wellbeing of women in male-factor infertility circumstances. These determinants particularly threaten the already challenged quality of life of the women in the lower socioeconomic groups.

This scoping review reveals a dearth of information on the effect of the socioeconomic status on the wellbeing of women in male-factor infertility marital relationships. The results emphasize social class and affordability as central to accessibility of NARTs. Couples’ socioeconomic statuses facilitate some choices and constrain others, with differences in wellbeing outcomes, capacities for relationship maintenance, and the development of adaptive strategies.

Emphasis should be on ensuring women’s socioeconomic empowerment especially in the LMICs. Their empowerment economically and educationally will mitigate the impact of male-factor infertility and other social ills on the women by affording them the requisite knowledge and exposure for developing the right help-seeking behaviours.

## Data Availability

The data sources have been included in the reference section of the submitted manuscript and are accessible.

## Ethical Considerations

- No conflicts of interest
- Only duly referenced published articles were reviewed
- No direct reference to any particular individual.

^**^S1 Text – Supporting Info – PRISMA-ScR Checklist

## Notes

### Competing Interest Statement

The authors have declared no competing interest.

### Funding Statement

The author(s) received no specific funding for this work.

